# A framework for quantifying the coupling between brain connectivity and heartbeat dynamics: Insights into the disrupted network physiology in Parkinson’s disease

**DOI:** 10.1101/2023.07.20.23292942

**Authors:** Diego Candia-Rivera, Marie Vidailhet, Mario Chavez, Fabrizio de Vico Fallani

**Affiliations:** Sorbonne Université, Paris Brain Institute (ICM), CNRS UMR7225, INRIA Paris (Nerv Team), INSERM U1127, AP-HP Hôpital Pitié-Salpêtrière, F75013, Paris, France

**Keywords:** Parkinson’s disease, Brain-heart interaction, Network physiology, Dopamine, Interoception

## Abstract

Parkinson’s disease (PD) often shows disrupted brain connectivity and autonomic dysfunctions, progressing alongside with motor and cognitive decline. Recently, PD has been linked to a reduced sensitivity to cardiac inputs, i.e., cardiac interoception. Altogether, those signs suggest that PD causes an altered brain-heart connection whose mechanisms remain unclear. Our study aimed to explore the large-scale network disruptions and the neurophysiology of disrupted interoceptive mechanisms in PD. We focused on examining the alterations in brain-heart coupling in PD and their potential connection to motor symptoms. We developed a proof-of-concept method to quantify relationships between the co-fluctuations of brain connectivity and cardiac sympathetic and parasympathetic activities. We quantified the brain-heart couplings from EEG and ECG recordings from PD patients on and off dopaminergic medication, as well as in healthy individuals at rest. Our results show that the couplings of fluctuating alpha and gamma connectivity with cardiac sympathetic dynamics are reduced in PD patients, as compared to healthy individuals. Furthermore, we show that PD patients under dopamine medication recover part of the brain-heart coupling, in proportion with the reduced motor symptoms. Our proposal offers a promising approach to unveil the physiopathology of PD and promoting the development of new evaluation methods for the early stages of the disease.

## Introduction

The understanding about the physiopathology and clinical phenotype of Parkinson’s disease (PD) remains limited. PD is known to affect motor function, but non-motor symptoms such as autonomic dysfunction have a significant impact on patients’ quality of life [Schapira et al., 2017]. Autonomic dysfunction can involve various bodily systems, including gastrointestinal, cardiovascular, urinary, erectile, thermoregulatory, and pupil contraction systems [Jain, 2011; Sharabi et al., 2021]. PD may also disrupt the awareness of one’s own heartbeats, as measured from cardiac interoception tasks [Hazelton et al., 2023; Ricciardi et al., 2016; Salamone et al., 2021; Santangelo et al., 2018], suggesting a disruption in the communication between the brain and the heart. Traditionally, cardiac interoception research relied on patients sensing their heartbeats through bodily sensations [Garfinkel et al., 2015]. However, recent advances have highlighted the value of objective markers of brain-heart interactions derived from physiological signals [Azzalini et al., 2019]. These markers represent the integrity and dynamics of the brain-heart communication, offering insights into the state of interoceptive pathways [Candia-Rivera, 2022]. This approach has proven valuable in diagnosing and predicting outcomes for brain-injured patients [Candia-Rivera and Machado, 2023; Hermann et al., 2024], demonstrating the potential of measuring brain-heart interactions to assess physiological states.

The study of PD physiopathology has tried to identify various patterns of brain activity to characterize the condition [Conti et al., 2022; Jackson et al., 2019; Leviashvili et al., 2022; Swann et al., 2015]. Because PD is a condition that can affect multiple parts of the nervous system, rather than being exclusively a focal brain region pathology [Gratton et al., 2019; Wang et al., 2021]. These physiological changes may not necessarily serve as definitive hallmarks for characterizing the disease, which is rooted in our limited understanding of their underlying mechanisms [Palma and Kaufmann, 2014]. Notably, research has shown that the damage occurring in multiple parts of the nervous system impacts global brain dynamics [Hammond et al., 2007]. However, understanding how this neural damage disrupts normal oscillatory functioning, leading to the disruption of motor functions, remains unclear [Candia-Rivera et al., 2024; Silberstein et al., 2005; Weinberger et al., 2006]. Studies employing various approaches have revealed that PD causes abnormal connectivity throughout various levels of the basal ganglia–cortical loop [Rivlin-Etzion et al., 2006], which may better explain motor deficits. The inhibition of this abnormal connectivity has given insights into diverse therapeutic strategies in mitigating motor impairment in Parkinson’s disease patients [Kühn et al., 2008; Wingeier et al., 2006].

In this study, we propose examining PD through the investigation of large-scale network disruptions, including those affecting the brain-heart communication. Given existing evidence suggesting that PD induces alterations in global brain networks [Hammond et al., 2007] alongside autonomic dysfunctions [Candia-Rivera et al., 2024; Sharabi et al., 2021], our hypothesis posits that a more comprehensive characterization can be achieved by exploring how the disease alters the brain-heart connection and understanding its relationship with dopamine and disrupted motor function. In this line, existing evidence has revealed that brain-heart interplay has a close relationship with motor excitability and associated behavioral responses [Agrimi et al., 2023; Al et al., 2023; Chen et al., 2023; Larra et al., 2020; Palser et al., 2021; Rae et al., 2018; Ren et al., 2022]. However, in the PD realm, limited evidence exists on their brain-heart interplay, which uniquely refers to changes in the central control of cardiac dynamics are linked with the severity of autonomic dysfunctions [Iniguez et al., 2022].

We hypothesized that PD alters the interplay between brain connectivity and cardiac dynamics, and that these alterations may be associated to PD symptoms beyond dysautonomia, such as motor outcomes, as a result of the disruption of the mechanisms in charge of shaping brain network dynamics [Shine, 2019]. This framework goes beyond state-of-the-art approaches for estimating brain-heart interplay [Candia-Rivera et al., 2021], which typically rely on gathering interactions between heartbeats and a single brain region. To do this, we propose a new framework for quantifying the relationship between brain connectivity and cardiac sympathetic and parasympathetic activities. Our study includes EEG and ECG from 16 healthy participants and 15 patients with mild to moderate PD stage [George et al., 2013], that underwent motor evaluation. PD patients were measured on and off dopaminergic therapy to further assess the impact of medication on motor symptoms and brain-heart interactions.

## Materials and methods

### Dataset description

The dataset [George et al., 2013; Rockhill et al., 2021] includes 15 PD patients (7 males and 8 females, median age = 63± 8 years) and 16 healthy participants (HS, 7 males and 9 females, median age = 60.5±8 years). The median disease duration is 3±2 years (range 1 to 12 years). PD patients were diagnosed by a movement disorder specialist at Scripps clinic in La Jolla, California. The patients were assessed using the Unified Parkinson’s Disease Rating Scale (UPDRS), section III, to evaluate the motor symptoms [Ramaker et al., 2002], whose scores are presented in Table I.

**Table I.** Dataset demographic information and clinical assessments.

| Participant id | Age range (years) | Sex | Disease duration (years) | UPDRS III (on) | UPDRS III (off) |
| --- | --- | --- | --- | --- | --- |
| HS 1 | 51-55 | f | n/a |  |  |
| HS 2 | 51-55 | f | n/a |  |  |
| HS 3 | 51-55 | f | n/a |  |  |
| HS 4 | 51-55 | f | n/a |  |  |
| HS 5 | 71-75 | f | n/a |  |  |
| HS 6 | 56-60 | f | n/a |  |  |
| HS 7 | 56-60 | m | n/a |  |  |
| HS 8 | 66-70 | m | n/a |  |  |
| HS 9 | 70-75 | m | n/a |  |  |
| HS 10 | 61-65 | f | n/a |  |  |
| HS 11 | 71-75 | f | n/a |  |  |
| HS 12 | 56-60 | f | n/a |  |  |
| HS 13 | 66-70 | m | n/a |  |  |
| HS 14 | 81-85 | m | n/a |  |  |
| HS 15 | 56-60 | m | n/a |  |  |
| HS 16 | 71-75 | m | n/a |  |  |
| PD 1 | 51-55 | f | 9 | 33 | 43 |
| PD 2 | 66-70 | f | 2 | 22 | 28 |
| PD 3 | 61-65 | f | 8 | 34 | 40 |
| PD 4 | 51-55 | f | 12 | 16 | 32 |
| PD 5 | 71-75 | f | 1 | 20 | 20 |
| PD 6 | 71-75 | m | 1 | 44 | 47 |
| PD 7 | 61-65 | m | 2 | 54 | 49 |
| PD 8 | 61-65 | m | 2 | 31 | 38 |
| PD 9 | 71-75 | m | 2 | 27 | 32 |
| PD 10 | 51-55 | m | 2 | 30 | 30 |
| PD 11 | 66-70 | f | 6 | 30 | 38 |
| PD 12 | 48-50 | m | 6 | 42 | 58 |
| PD 13 | 66-70 | f | 3 | 36 | 44 |
| PD 14 | 71-75 | f | 3 | 25 | 42 |
| PD 15 | 61-65 | m | 9 | 46 | 48 |

PD: Parkinson's disease, HS: Healthy state, f: Female, m: Male, UPDRS: Unified Parkinson's Disease Rating Scale, n/a: It does not apply.

Participants provided written consent in accordance with the Institutional Review Board of the University of California, San Diego, and the Declaration of Helsinki. Details on the demographic information of each participant are available in the original studies from this cohort [George et al., 2013; Rockhill et al., 2021].

PD patients’ data were collected under on- and off-medication. On- and off-medication conditions were collected on different days with a counterbalanced order. For the on-medication recordings, patients continued their typical medication regimen. For the off-medication state, patients discontinued medication use at least 12 h before the session.

EEG data were acquired using a 32-channel BioSemi ActiveTwo system, together with a one-lead ECG, sampled at 512 Hz at rest for approximately 3 min.

### EEG processing

EEG data were pre-processed using MATLAB R2022b and Fieldtrip Toolbox [Oostenveld et al., 2011]. Data were bandpass filtered with a fourth-order Butterworth filter, between 0.5 and 45 Hz. To mitigate substantial movement artifacts, we employed a wavelet-enhanced independent component analysis (ICA) for the efficient removal of these artifacts from individual components [Gabard-Durnam et al., 2018]. Subsequently, we reconstructed EEG signals for further analysis. A second round of ICA was conducted to specifically identify components associated with eye movements and cardiac-field artifacts, which were then systematically set to zero. To this end, one lead ECG was included as an additional input to the ICA to enhance the process of finding cardiac artifacts. Once the ICA components with eye movements and cardiac artifacts were visually identified, they were set to zero to reconstruct the EEG series. The results of this step were eye-movements and cardiac-artifact-free EEG data. Channels were re-referenced using a common average [Candia-Rivera et al., 2021].

### ECG processing

ECG time series were bandpass filtered using a fourth-order Butterworth filter, between 0.5 and 45 Hz. The R-peaks from the QRS waves were identified with an automatized process, followed by a visual inspection of misdetections. The procedure was based on a template-based method for detecting R-peaks [Candia-Rivera et al., 2021]. All the detected peaks were visually inspected over the original ECG, along with the inter-beat intervals histogram. Manual corrections of misdetections were performed if needed. The mean RR intervals, together with cardiac sympathetic and parasympathetic activities were computed to compare HS vs PD, and on vs off dopamine medication conditions.

### Computation of cardiac sympathetic and parasympathetic indices

The cardiac sympathetic and parasympathetic activities were estimated through a method based on the time-varying geometry of the interbeat interval (IBI) Poincaré plot [Candia-Rivera, 2023]. Poincaré plot is a non-linear method to study heart rate variability and depicts the fluctuations on the duration of consecutive IBIs [Brennan et al., 2001]. The features quantified from Poincaré plot are the SD_1_ and SD_2_, the ratios of the ellipse formed from consecutive changes in IBIs, representing the short- and long-term fluctuations of heart rate variability, respectively [Sassi et al., 2015].

The ellipse ratios for the whole experimental condition SD_01_ and SD_02_ are computed as follows:

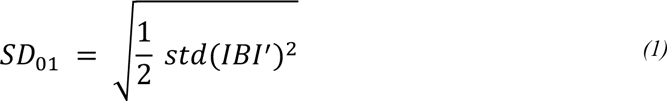

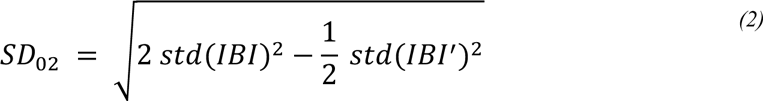

where IBI^’^ is the derivative of IBI and std() refers to the standard deviation.

The fluctuations of the ellipse ratios are computed with a sliding-time window, as shown in Eq. *(3)* and *(4)*:

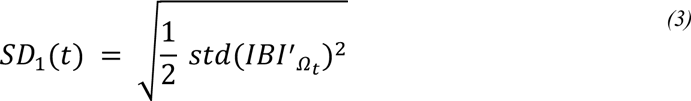

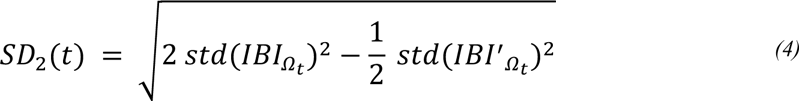

where Ω_t_: t – T ≤ t_i_ ≤ t, in this study T is fixed in 15 seconds.

The Cardiac Parasympathetic Index (CPI) and Cardiac Sympathetic Index (CSI) are computed as follows:

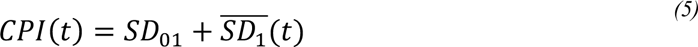

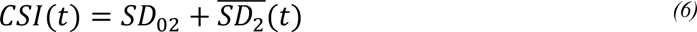

where 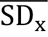 is the demeaned SD_x_

For a comprehensive description of the method, see [Candia-Rivera, 2023].

### EEG connectivity fluctuations

The EEG spectrogram was computed using the short-time Fourier transform with a Hanning taper. Calculations were performed through a sliding time window of 2 seconds with a 50% overlap, resulting in a spectrogram resolution of 1 second and 0.5 Hz. Time series were integrated within three frequency bands (alpha: 8-12 Hz, beta: 12-30 Hz, gamma: 30-45 Hz). Those definitions were based on previous EEG connectivity findings in PD, e.g., [Conti et al., 2022]. It is important to note that the definition of frequency bands, as well as the overlap between them, exhibits some variability in the literature. Therefore, a careful consideration should be given to these definitions.

Since connectivity measures were assessed at the scalp level, we performed symmetry tests to ensure that participant groups did not significantly differ in asymmetry-symmetry balance, which could arise due to volume conduction artifacts. To achieve this, we calculated the Asymmetry-Symmetry Ratio (ASR) developed by Haufe and colleagues [Haufe et al., 2012]. This ratio offers an indication of asymmetry-symmetry balance based on covariance matrices across EEG channels. ASR computations were conducted for the alpha, beta, and gamma bands. We then utilized Wilcoxon-Mann-Whitney tests to assess whether ASR varied across the three conditions: healthy state, Parkinson’s disease on dopamine medication, and off dopamine medication.

The directed time-varying connectivity between power series of two EEG channels was quantified using an adaptative Markov process [Al-Nashash et al., 2004]. The algorithm consisted in a first-order autoregressive model, which was aimed for capturing the directed temporal dynamics of an EEG channel, by leveraging on the dependency of its past values and another EEG channel, represented as the external term in the model. The use of a first-order model provides a parsimonious description of the autocorrelation structure of the time series, with a good compromise on accuracy [Al-Nashash et al., 2004; Bai et al., 2001]. Moreover, the estimation of directed connectivity using an autoregressive model enables the quantification of causal relationships, rather than synchronization [Chiarion et al., 2023]. This is crucial for understanding the flow of information within the brain and identifying directional influences between specific channels.

The EEG series of the channel ch1 is represented as the sum of power series, as shown in Equation *(7)*, where *f* is the frequency (*f* = *f*_1_, …, *f_n_)*, *θ_f_* is the respective phase, and *a*_*f*_ is the power series within the frequency *f*. In this study, the frequency range considered was 1-45 Hz, with 0.5 Hz step. Then, the model estimates the directed connectivity at a specific frequency band (*F = {alpha, beta, gamma}*). Therefore, *a_F_* represents the power series integrated within the band *F*, which is modeled in a first order auto-regressive process. In Equation *(8)*, *a_F_* is modeled by estimating *A_F_* as the contribution of its past values, and by estimating *B_F_*, the contribution of the external term. The model is set to minimize the adjusted error *ε_F_,* using least squares. Then, the directed and time-varying connectivity is obtained from the adjusted coefficient from the external term *B_F_*, as shown in Equation *(9)*.

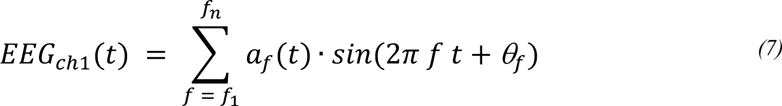

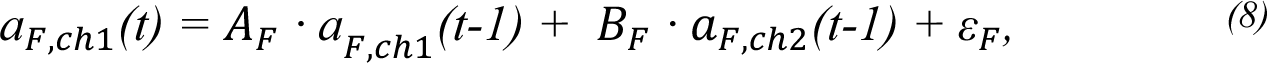

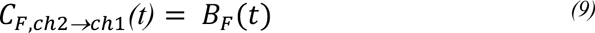

To validate the reliability of our connectivity modeling, we conducted control analyses to ensure it was well fitted. We employed the normalized Akaike Information Criterion (nAIC) [Akaike, 1974] within the healthy participant subset, focusing on the alpha band. Specifically, we compared the goodness of fit of order 1 models against order 2 and 3 models.

For each sliding time window, EEG channel pair, and subject, we computed nAIC values for all models. These values were then grand-averaged across time and participants to ensure that the difference in nAIC between the order 1 model and the order 2 and 3 models did not exceed two [Burnham and Anderson, 2004].

Finally, we used a Kolmogorov-Smirnov test to verify if nAIC values from order 1 models across participants come from the same distribution as those from order 2 and 3 models.

### Brain-heart coupling estimation

As depicted in Figure 1, brain-heart coupling was quantified by considering the relationships between brain connectivity fluctuations and cardiac sympathetic-parasympathetic indices. The brain-heart coupling was assessed using Maximal Information Coefficient (MIC). MIC is a method that quantifies the coupling between two time series [Reshef et al., 2011]. MIC evaluates similarities between different segments separately at an adapted time scale that maximizes the mutual information, with a final measure that wraps the similarities across the whole time-course. The Equations *(10)* and *(11)* show the MIC computation between two time series X and Y. The mutual information *I_g_* is computed to different grid combinations *g* ∈ *G_xy_*. The mutual information values are normalized by the minimum joint entropy log_2_ min{*n_x_*, *n_y_*}, resulting in an index in the range 0-1. Finally, the quantified coupling between X and Y corresponds to the normalized mutual information resulting from the grid that maximizes the MIC value.

**Figure 1.**
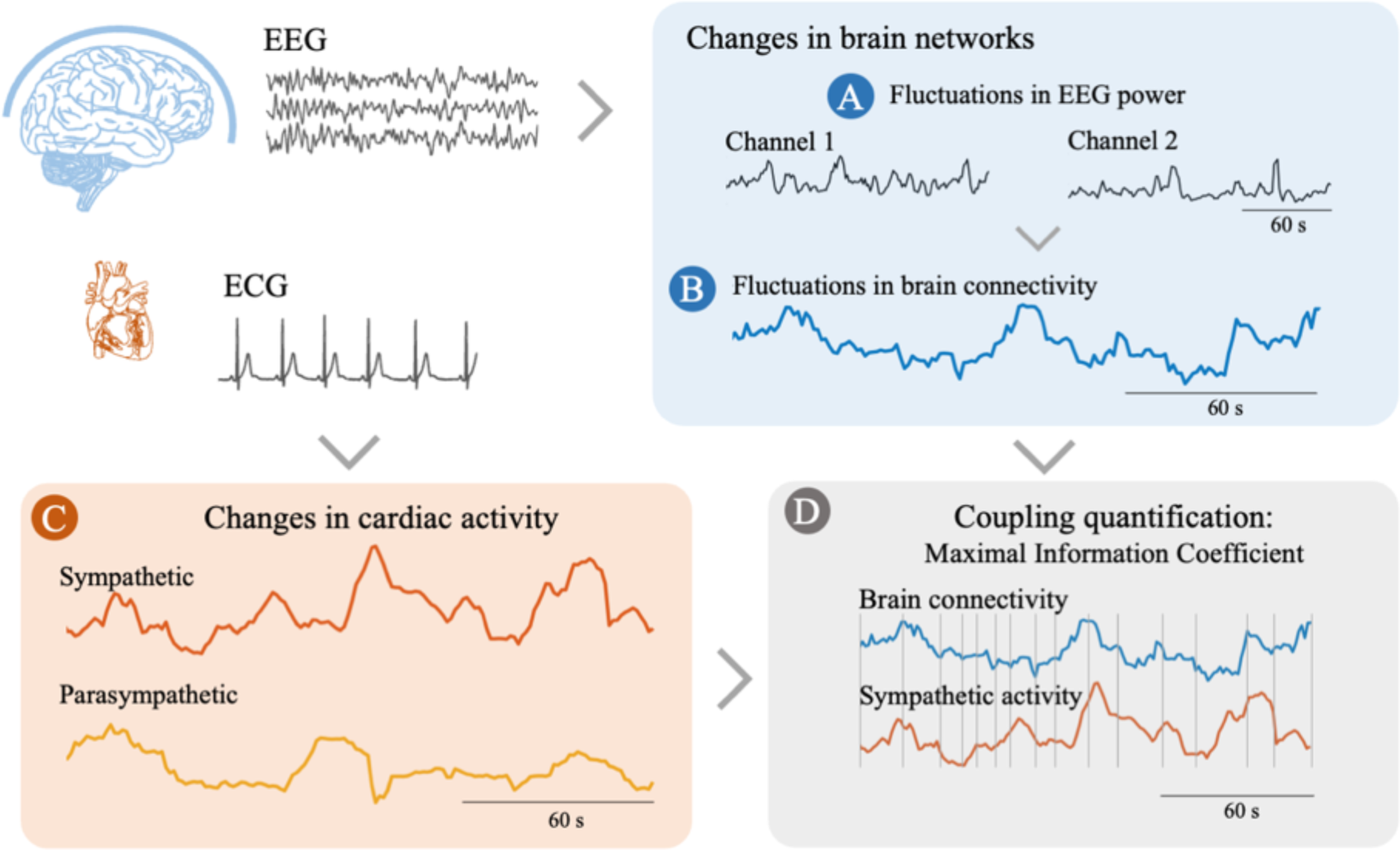
Methodological pipeline. (A) Computation of time-varying EEG power at different frequency bands (α, β, γ) and (B) the estimation of time-varying connectivity between two EEG channels. (C) Computation of the heart rate variability series from ECG and the estimation of cardiac sympathetic-parasympathetic activity. (D) Brain connectivity-cardiac coupling estimation by computing the Maximal Information Coefficient (MIC). The coupling quantification is achieved by assessing the similarities between two time series, regardless of the curvature of the signals. The MIC method evaluates similarities between distinct segments individually, using an adjusted grid as depicted in the figure. The overall measure combines the similarities observed throughout the entire time-course.

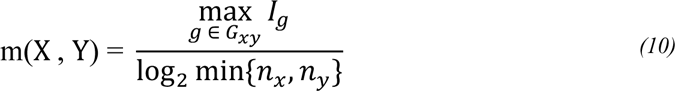

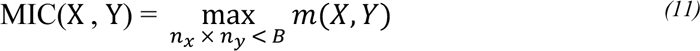

where B = *N*^0.6^, and N is the dimension of the signals [Reshef et al., 2011]. The source code implementing MIC is available online at https://github.com/minepy.

### Heartbeat-evoked responses analysis

Heartbeat-evoked responses (HERs) were defined by averaging time-locked EEG epochs with respect to R-peaks, from 0 to 500 ms [Park and Blanke, 2019a]. For HERs computation, the EEG epochs selection followed two rules: (i) epochs maximum absolute amplitude < 300 μV on any EEG channel, and (ii) the next heartbeat occurred at a latency later than 500 ms.

### Statistical analysis

MIC values were compared between groups: healthy state vs Parkinson’s disease on dopamine, healthy state vs Parkinson’s disease off dopamine, and Parkinson’s disease on vs off dopamine. Statistical comparisons were based on Wilcoxon-Mann Whitney signed rank and rank sum tests, for paired and unpaired comparisons, respectively. P-values were corrected for multiple comparisons by using cluster-permutation analyses.

Cluster permutation analysis was performed with the objective of identifying paired and unpaired differences in brain activity, applied in this study to different measures quantifying brain-heart coupling. This approach addresses the challenge of multiple comparisons by detecting clusters of adjacent data points where differences occur and using permutation testing to control for false positives [Maris and Oostenveld, 2007]. Clustered effects were revealed using a non-parametric version of cluster permutation analysis [Candia-Rivera and Valenza, 2022]. Cluster permutation analysis was applied to HERs and the MIC values computed between the directed connectivity and cardiac sympathetic/parasympathetic activity.

The algorithm to identify and test the significance of cluster in the data included a preliminary mask definition, identification of candidate clusters and the computation of cluster statistics with Monte Carlo’s p-value correction:

1. The preliminary mask was defined from squared matrices containing the MIC values, indicative of brain-heart coupling for each pair of channels. The matrices were composed by 992 values (32 x 32 channels, minus the diagonal). Wilcoxon-Mann Whitney tests were applied 992 times, as depicted in Figure 2A. The preliminary mask was then defined by the threshold on the p-value from the Wilcoxon-Mann Whitney tests, defined at α=0.05. Note that in this study, matrices are not symmetrical because directed connectivity measures between every pair of channels were computed.
2. Candidate clusters were identified based on neighboring points within the preliminary mask. As shown in Figure 2B, for a given channel pair channel_1_→channel_2_, the neighboring connections include both the connections from channel_1_ to the neighbors of channel_2_ and the connections from the neighbors channel_1_ to the neighbors of channel_2_. The default Biosemi neighborhood definition for 32 channels was used, and a minimum cluster size of 5 neighbors was imposed to proceed.
3. Cluster statistics were computed in each preliminary cluster identified from the previous step. The MIC values from all the points pertaining to one candidate cluster were averaged and tested against 10,000 random partitions. The proportion of random partitions that resulted in a lower p-value than the observed one was considered as the Monte Carlo p-value. The significance of the Monte Carlo p-values was set at α=0.05. The cluster statistic considered is the Wilcoxon-Mann Whitney’s absolute maximum Z-value obtained from all the points pertaining to the cluster. As depicted in Figure 2C, the process begins by identifying from all possible channel pairs those with a priori significance in the preliminary mask. Subsequently, through cluster identification, individual networks are discerned from the initial set of channel pairs. The visualization of the brain networks was performed using Vizaj [Rolland and Fallani, 2023].

**Figure 2.**
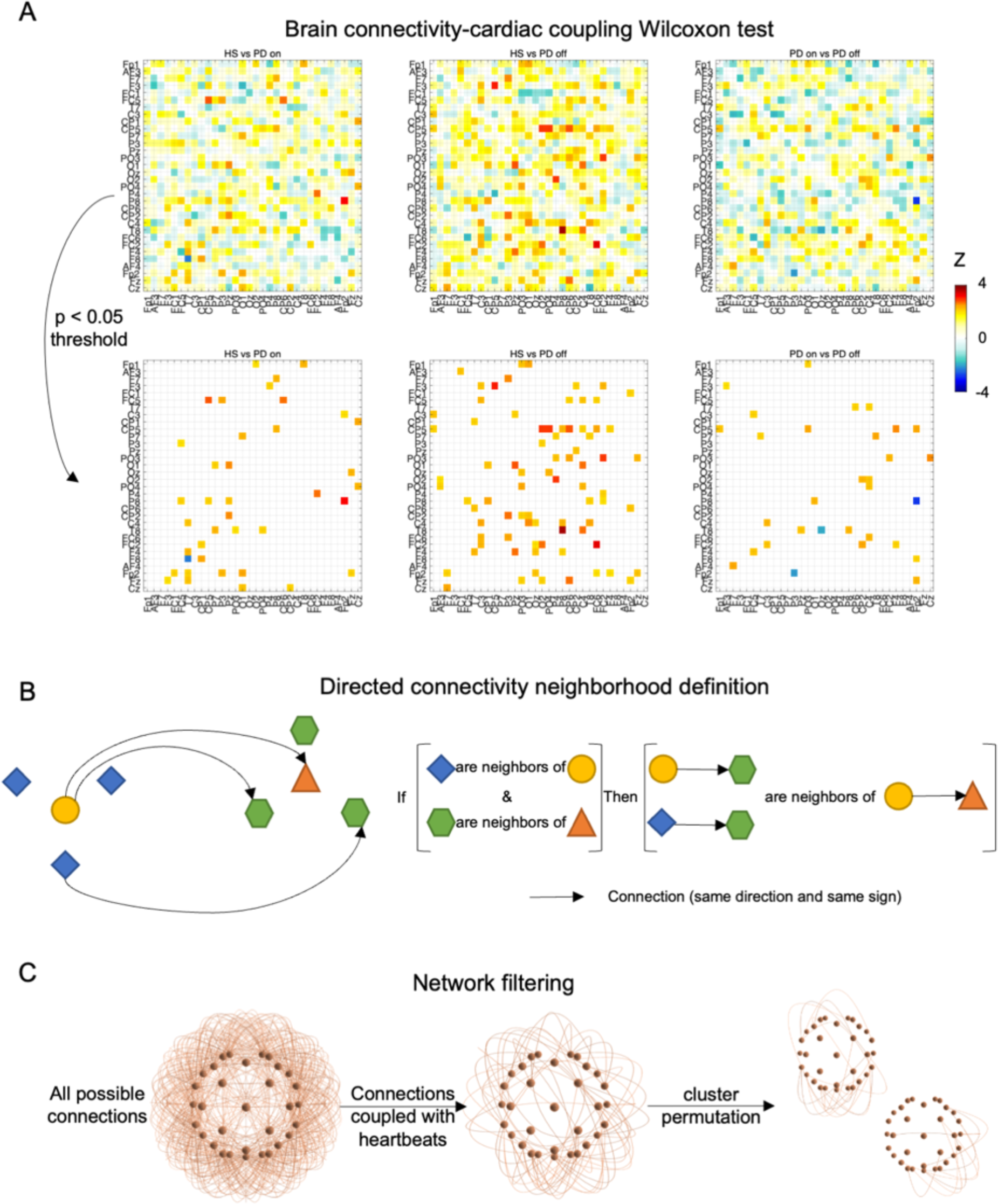
Network cluster permutation pipeline. (A) The connections that resulted in a p-value lower than the defined critical alpha are retained for constructing a preliminary mask for further analysis. (B) Neighboring connections are grouped by following the neighboring rule displayed. (C) Cluster statistics are computed for all the averaged connections that belong to the cluster and corrected for 10,000 permutations.

We further analyzed the identified networks showing differences on the brain-heart coupling by quantifying the relationship of the mean brain-heart coupling values with UPDRS-III scores using Spearman Correlation. Significance of the correlation analysis was defined by the Bonferroni rule α = 0.05/N, with N equal to the number of networks identified.

### Resource availability

The data is part of a publicly available dataset “UC San Diego Resting State EEG Data from Patients with Parkinson’s Disease”, gathered from OpenNeuro.org the 21^st^ of November of 2022 [Appelhoff et al., 2019; Pernet et al., 2019; Rockhill et al., 2021].

The utilized code come from different toolboxes for MATLAB. The functions for the computation of cardiac sympathetic and parasympathetic indices [Candia-Rivera, 2023] are available at https://github.com/diegocandiar/robust_hrv. The functions for the computation of time-varying connectivity and brain-heart coupling are available at https://github.com/diegocandiar/heart_brain_conn. The functions for the computation of MIC values [Reshef et al., 2011] are available at https://github.com/minepy. The functions to perform cluster permutation analyses [Candia-Rivera and Valenza, 2022] are available at https://github.com/diegocandiar/eeg_cluster_wilcoxon. The data analysis was performed using Fieldtrip toolbox [Oostenveld et al., 2011], available at https://github.com/fieldtrip/fieldtrip

## Results

We computed the coupling between brain connectivity and cardiac sympathetic and parasympathetic activities by considering linear and nonlinear associations between the brain and the heartbeat-derived time series. Initially, we computed EEG power series within the alpha, beta, and gamma bands. Given that our pipeline includes scalp-level connectivity analysis, we ensured that the conditions being compared did not significantly vary in their asymmetry-symmetry balance, which could stem from volume conduction artifacts. We observed only a slight difference in the gamma band (refer to Supplementary Material, Table S1). Subsequently, we calculated directed and time-resolved connectivity measures using a Markovian process, validating their goodness of fit through nAIC analyses (refer to Supplementary Material, Figures S1 and S2).

We compared the brain-heart couplings between healthy individuals and patients with PD, on and off dopaminergic therapy. We observed significant variations in the relationship between the brain connectivity and heartbeat dynamics among PD patients and healthy individuals. In healthy individuals, we observed a coupling between fluctuations in EEG connectivity and variations in cardiac dynamics. However, this coupling was weaker in PD patients, particularly in the relationship between slow fluctuations of heart rate variability (which are typically associated with sympathetic activity) and alpha and gamma connectivity.

We found differences when comparing the brain-heart coupling of healthy participants with that of PD patients who were not receiving dopaminergic therapy (PD off). Through cluster-based permutation tests, applied to the ensemble of EEG connectivity values coupled with heartbeat dynamics, we discovered that one network in the alpha band was significantly linked to cardiac sympathetic indices, whose coupling was reduced in PD (Figure 3A and B, cluster statistics HS vs PD off, p = 0.0002, Z = 2.9844, cluster size = 13). These findings indicate that the resting state neural dynamics in PD are disturbed, affecting the interactions between brain connectivity and heartbeat dynamics. Importantly, these differences can be identified using non-invasive methods, without requiring any form of stimulation.

**Figure 3.**
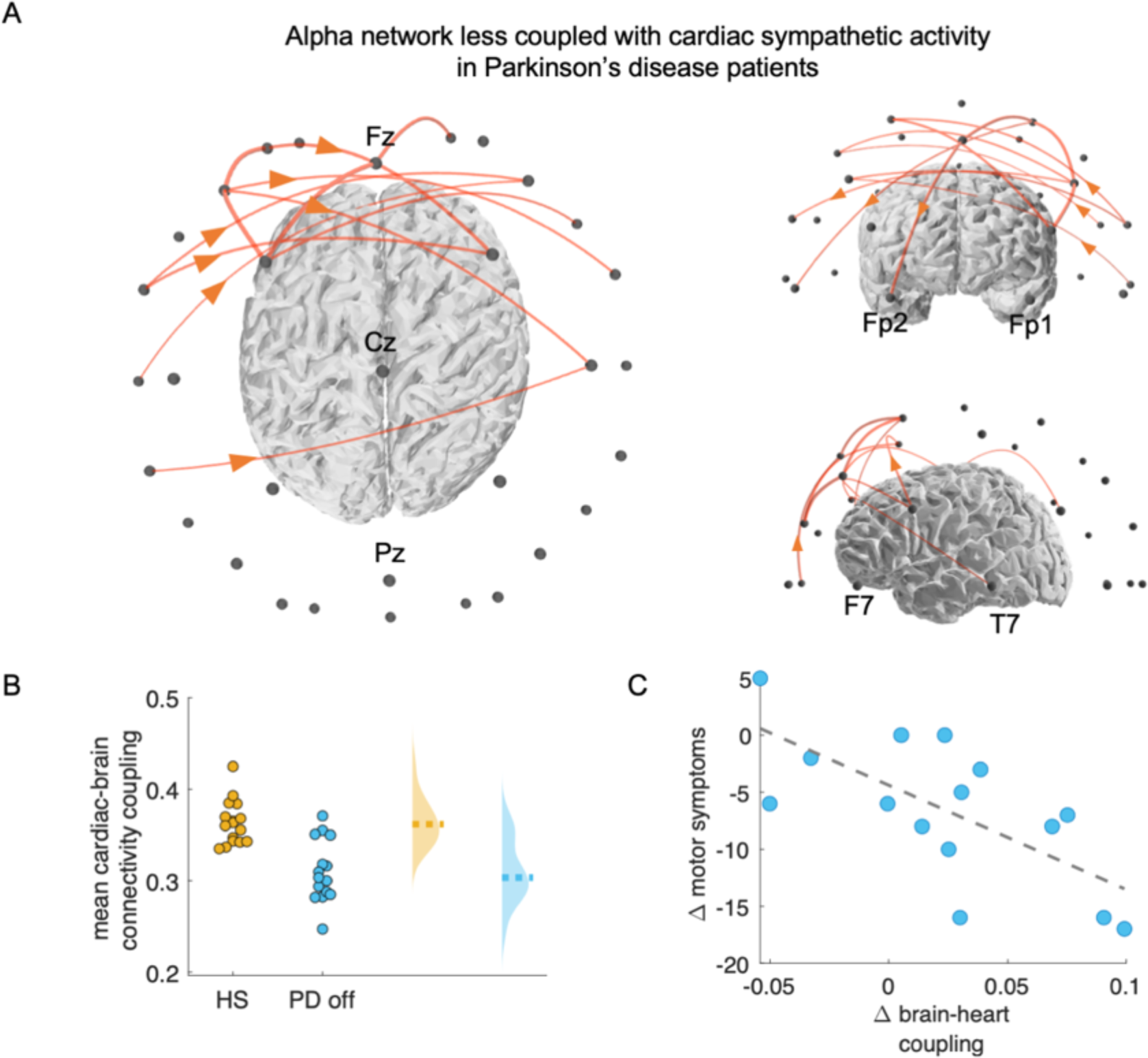
Significant alpha network that correlated with cardiac sympathetic indices. (A) The network distinguishing healthy participants from PD patients off dopaminergic therapy. (B) Distribution of the mean brain-heart coupling. The dashed lines indicate the group medians. (C) Correlation between the changes in the brain-heart coupling (ι1 brain-heart coupling, i.e., on minus off) and the changes in motor symptoms (motor section of the United Parkinson’s Disease Rating Scale—where a lower score means better motor outcome). All values are in arbitrary units.

Dopaminergic medication significantly improved motor symptoms, as measured by the motor section of the Unified Parkinson’s Disease Rating Scale–UPDRS III, as performed in a paired Wilcoxon test (Z = 2.9388, p = 0.0033). Furthermore, our results suggest that dopaminergic therapy is associated with the increased brain-heart coupling in patients with PD. We found a correlation between those changes in brain-heart coupling and the changes in the motor evaluation in PD patients (as evaluated in the motor section of the Unified Parkinson’s Disease Rating Scale–UPDRS III [Ramaker et al., 2002]). Specifically, the significant correlation between the improvement in motor symptoms and brain-heart coupling was found in the alpha band (Figure 3C, Spearman correlation, R = 0.6470, p = 0.0091). This suggests that measures of brain-heart coupling are sensitive to the physiological changes induced by dopaminergic therapy in PD patients.

Additionally, we found two networks in the gamma band that were also linked to the estimation of cardiac sympathetic indices as well. These two networks were located in the parieto-frontal (Cluster statistics HS vs PD off, p < 0.0001, Z = 2.9844, cluster size = 21) and parieto-temporal regions (Cluster statistics HS vs PD off, p < 0.0001, Z = 2.7472, cluster size = 11), as shown in Figure 4 (please note that by reducing the gamma band leads to the merging of these two networks, see Supplementary Material Figure S3). These couplings showed no correlation with changes in motor symptoms in either gamma network 1 (Spearman correlation, R = 0.4050, p = 0.1342), or in gamma network 2 (Spearman correlation, R = 0.4444, p = 0.0969).

**Figure 4.**
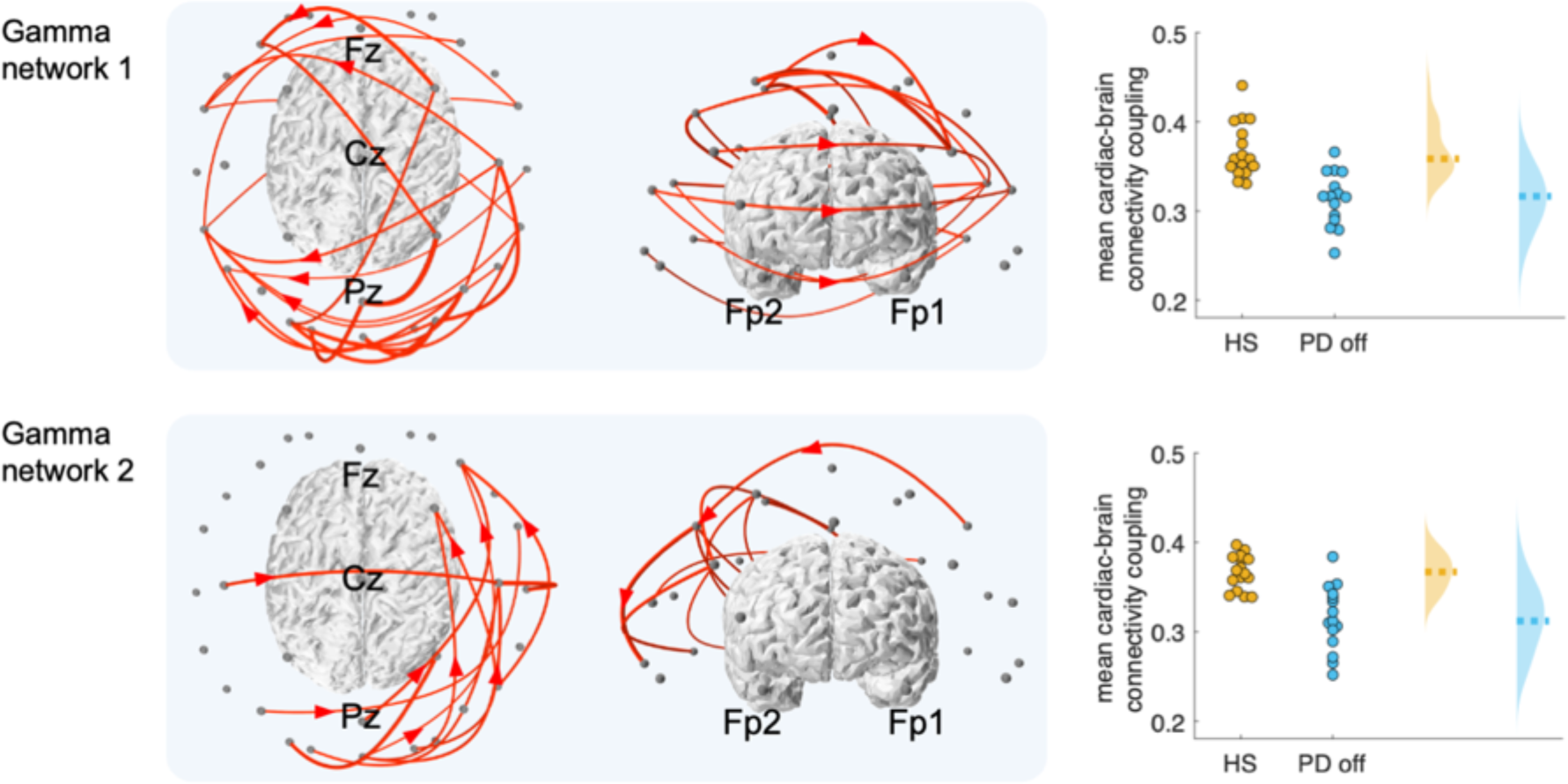
Significant gamma networks that correlated with cardiac sympathetic indices, distinguishing healthy participants from PD patients off dopaminergic therapy. The distributions of the mean brain-heart coupling values are displayed for each network. The dashed lines indicate the group medians. All values are in arbitrary units.

As shown in Table II, we found non-significant differences when comparing the mean connectivity values in the identified alpha network, gamma network 1 and gamma network 2, and cardiac sympathetic and parasympathetic indices separately. A moderate difference emerged only when comparing the RR interval in HS vs PD on dopamine medication. However, this appears not to be influencing the findings in brain-heart coupling measures, which mainly distinguished between HS and PD off dopamine medication.

**Table II.** Wilcoxon-Mann Whitney tests on the comparisons of the mean connectivity values in the identified alpha and gamma networks, and in the RR interval, cardiac sympathetic and parasympathetic indices. Comparisons were performed between healthy state (HS) and Parkinson’s disease (PD) patients, on and off dopamine.

|  | Alpha network | Gamma network 1 | Gamma network 2 | RR interval | Cardiac Sympathetic index | Cardiac Parasympathetic index |
| --- | --- | --- | --- | --- | --- | --- |
| HS vs PD on | $Z=0.8103$,<br>$p=0.4177$ | $Z=0$, $p=1$ | $Z=1.6800$,<br>$p=0.0930$ | $Z=2.3915$,<br>$p=0.0168$ | $Z=-1.0870$,<br>$p=0.2770$ | $Z=-0.2174$,<br>$p=0.8279$ |
| HS vs PD off | $Z=-0.5336$,<br>$p=0.5936$ | $Z=-0.9684$,<br>$p=0.3328$ | $Z=1.5614$,<br>$p=0.1184$ | $Z=1.9567$,<br>$p=0.0504$ | $Z=0.6522$,<br>$p=0.5143$ | $Z=0.8499$,<br>$p=0.3954$ |
| PD on vs PD off | $Z=-1.3063$,<br>$p=0.1914$ | $Z=-1.0223$,<br>$p=0.3066$ | $Z=0.1704$,<br>$p=0.8647$ | $Z=-0.1244$,<br>$p=0.9010$ | $Z=0.3976$,<br>$p=0.6909$ | $Z=-0.5680$,<br>$p=0.5701$ |

Furthermore, we examined the relationship between the brain and heartbeats by analyzing heartbeat-evoked responses (HERs), acknowledged markers of the central processing of cardiac inputs [Park and Blanke, 2019b]. HERs were gathered from the average of EEG epochs synchronized with the cardiac cycle. We compared HERs in healthy individuals to those with PD, both on and off dopaminergic therapy, and compared HERs in PD patients on and off dopamine therapy. Our findings revealed distinct HER patterns when comparing PD patients on and off dopaminergic therapy (Cluster statistics PD on vs PD off. Positive clusters: p_1_ = 0.0008, Z_1_ = 3.2942; p_2_ = 0.0068, Z_2_ = 3.0102. Negative cluster: p = 0.0037, Z = 2.8966), as shown in Figure 5. However, there was only a slight difference between the two conditions, suggesting that higher-order brain-heart interaction analysis, such as the coupling between cardiac and brain networks may be a more suitable approach for characterizing PD.

**Figure 5.**
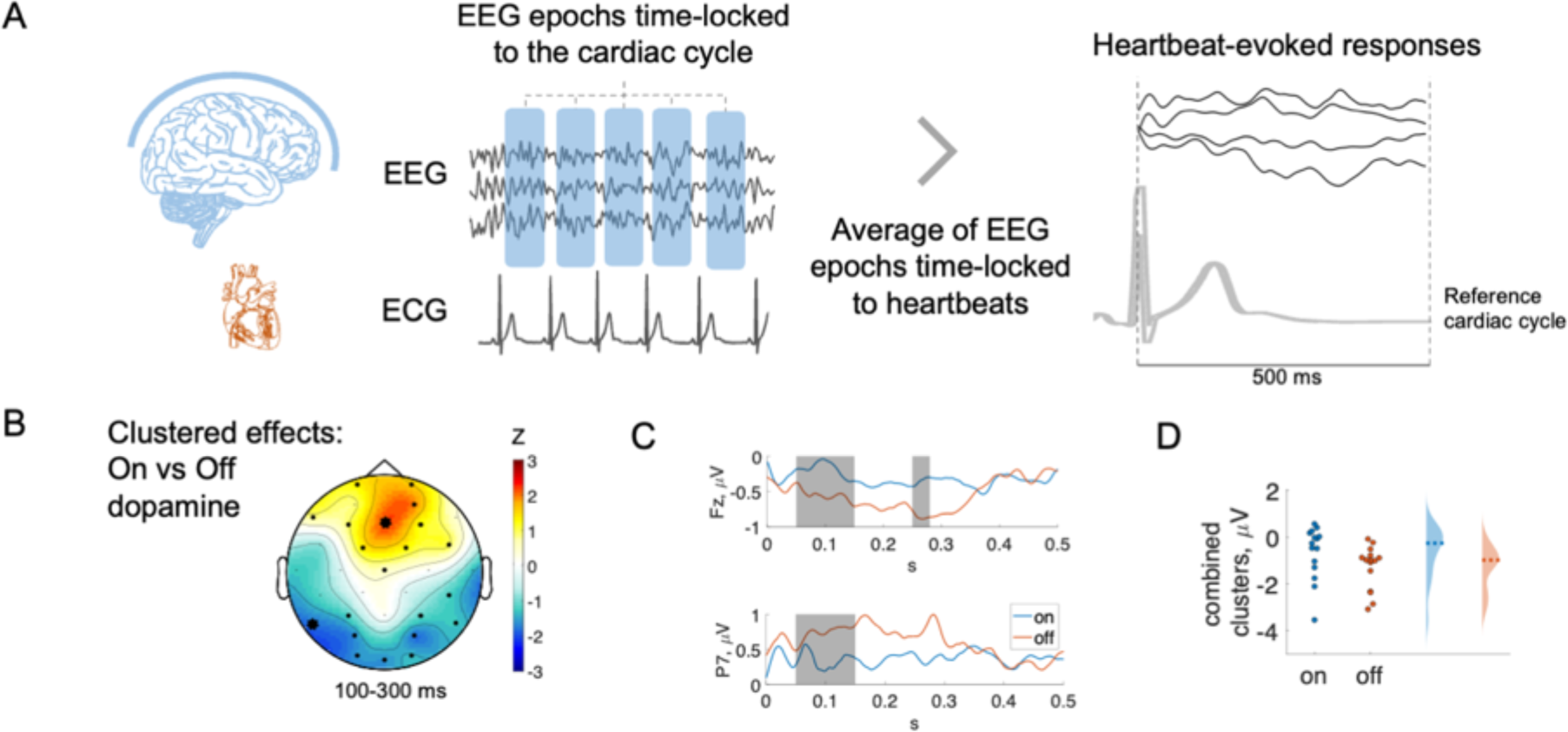
Heartbeat-evoked responses (HERs). (A) Pipeline to compute HERs. (B) Clustered effects found when comparing PD on vs PD off. Thick channels show clustered effects, and the color bar indicates the Z-value obtained from the paired Wilcoxon test. (C) Group median time course of the thick electrodes shown in (A). (D) Combined clustered effects. PD on: Parkinson’s disease on dopamine, PD off: Parkinson’s disease off dopamine.

## Discussion

The physiological basis of disrupted cardiac interoceptive pathways in PD, as assessed by brain-heart interactions, has not been significantly explored to date. We found that the couplings of fluctuating alpha and gamma connectivity with cardiac sympathetic dynamics are reduced in PD patients off dopamine, as compared to healthy individuals. Furthermore, we show that PD patients on dopamine medication recover part of the brain-heart coupling, in proportion with the reduced motor symptoms.

Peripheral autonomic neurons can be affected in PD [Wakabayashi and Takahashi, 1997], leading to symptoms of dysautonomia ranging among cardiovascular, respiratory, gastrointestinal, urinary, erectile, thermoregulatory, and pupil contraction disorders [Jain, 2011; Sharabi et al., 2021]. The appearance of autonomic damage in PD has led to a search for specific abnormalities in autonomic function, e.g., heart rate variability [Devos et al., 2003; Haensch et al., 2009] and its synchronization with specific brain regions [Iniguez et al., 2022], that could predict the disease and their symptoms. The reliability of these biomarkers remains uncertain due to the lack of understanding regarding their underlying mechanisms [Palma and Kaufmann, 2014]. Furthermore, the strongest evidence indicates that autonomic markers may rather provide insights into the severity and prognosis of PD [Brisinda et al., 2021; De Pablo-Fernandez et al., 2017; Iniguez et al., 2022].

The exploration of the relationship between brain connectivity and cardiac dynamics in PD is motivated by the substantial evidence of abnormal brain connectivity and autonomic abnormalities found in PD patients. These findings may provide a link to the observed disruptions in interoception in these individuals [Hazelton et al., 2023; Ricciardi et al., 2016; Salamone et al., 2021; Santangelo et al., 2018]. Nigrostriatal fiber degeneration in PD disrupts the striato-cortical functional connectivity networks, leading to the known impairments in motor control [Ruppert et al., 2020]. However, in the early stages of PD, changes in brain metabolism occur in key nodes of motor and cognitive networks, which can lead to disruptions in the connectivity of several regions [Huang et al., 2007; Nigro et al., 2016]. This has motivated the study of PD in terms of network-level phenomena rather than focal pathology [Gratton et al., 2019; Wang et al., 2021]. We investigated how cortical connectivity and heart rate variability covary at resting state. Our study found notable differences between the coupling between cardiac dynamics and brain connectivity in patients with PD and healthy individuals. In healthy participants, we noticed that changes in time-varying EEG connectivity are linked to changes in cardiac dynamics. However, this coupling is reduced in PD patients, especially in the connection between slow fluctuations of heart rate variability (considered predominantly sympathetic) and alpha and gamma connectivity. When PD patients are under dopaminergic therapy, the brain-heart coupling changes, suggesting a close link between the changes triggered by dopamine replacement and brain-heart coupling measures. This may indicate that markers of brain-heart interactions can capture dopaminergic-dependent mechanisms that are disrupted in PD. Indeed, one of the pathways affected by PD is the locus coeruleus-noradrenaline pathway [Benarroch, 2009]. The disruptions in the locus coeruleus-noradrenaline pathway lead to changes in the slow fluctuations of heart rate variability, which are caused by changes in sympathetic activity resulting from variations in the noradrenaline release rate [Barcroft and Konzett, 1949].

The acknowledged multi-organ dysfunction found in PD indicate that the physiopathology involves the disruption of several interoceptive pathways [Jain, 2011; Sharabi et al., 2021], including cardiac sympathetic denervation caused by the loss of catecholamine innervation in the nigrostriatal system and in the sympathetic nervous system [Goldstein et al., 2000]. Indeed, catecholamines (noradrenaline and dopamine) and sympathetic pathways play a relevant role in the brain-heart communication mechanisms in healthy individuals [Lueckel et al., 2018]. Previous behavioral studies have found that some patients with PD have difficulty sensing their own heartbeats, as quantified from cardiac interoception tasks [Ricciardi et al., 2016; Santangelo et al., 2018]. This suggests that their brain-heart communication may be disrupted. In another study [Salamone et al., 2021] the authors found an improved emotion recognition when healthy individuals performed an emotion recognition task after completing a cardiac interoception task. However, this effect was not observed in patients with PD [Salamone et al., 2021]. Furthermore, early PD has reported atrophy of the insula, key structure in interoceptive processing [Claassen et al., 2016]. Interoceptive inputs have been recognized as playing an important role in perception within computational frameworks of predictive coding [Petzschner et al., 2021] and consciousness [Candia-Rivera, 2022], where dopamine is thought to be critical for processing interoceptive prediction errors [Seth et al., 2011; Spindler et al., 2021]. Numerous studies have shown that dopamine encodes learning and reward prediction [Fiorillo et al., 2003; Hollerman and Schultz, 1998; Mirenowicz and Schultz, 1994; Pessiglione et al., 2006], further supporting this idea. On account of the key role of dopamine-modulated mechanisms, it has been hypothesized that dopamine participates in adaptation processes in predictive coding [Corlett et al., 2010], which may extend to the role of dopamine in the regulation of the subjective experience of perception [Lou et al., 2011].

Our results may provide new insights for the understanding of the well-known abnormalities in brain connectivity of PD. For instance, PD patients show decreased connectivity in the supplementary motor area, dorsal lateral prefrontal cortex, and putamen, but increased connectivity in the cerebellum, primary motor cortex, and parietal cortex [Wu et al., 2009]. PD patients may have higher connectedness within the sensorimotor and visual networks [Göttlich et al., 2013], due to compensation or loss of mutual inhibition between brain networks. Dopamine medication can normalize some patterns of functional connectivity, but the recovery level may depend on disease severity [van Eimeren et al., 2009; Wu et al., 2009]. In our results we observed a decrease in the coupling of cardiac sympathetic activity with brain connectivity measured in the alpha and gamma bands. EEG studies have revealed significant changes in the alpha-gamma range in PD, with reduced connectivity in alpha-beta bands and increased connectivity in the gamma band [Conti et al., 2022], but also aberrant cortical synchronization in the beta band [Jackson et al., 2019; Swann et al., 2015]. It remains to be confirmed whether our results relate to the repeatedly reported changes in brain connectivity in PD, including subcortical structures.

Our study has limitations, such as a small sample size, the use of low-density EEG, short recordings, and only assessing patients at rest. Specifically, we refrained from conducting connectivity analysis on source-reconstructed data due to the limited density of available scalp recordings, a factor known to introduce biased estimations [Song et al., 2015]. Instead, our chosen sensor-level approach offers practical advantages, including computational efficiency and decreased susceptibility to inaccuracies associated with volume conduction and brain region specificity [Van de Steen et al., 2019]. While our method provides valuable insights into connectivity dynamics, caution should be considered when interpreting brain spatial details. Given the marginal differences observed in the symmetry of EEG data in the gamma band between healthy participants and those with Parkinson’s disease off dopamine, it should not be rejected the possibility that some of the links identified in the gamma networks could be attributed to volume conduction issues. Future studies, with a targeted focus on specific brain regions, may consider our framework by incorporating methods to mitigate volume conduction effects [Talebi et al., 2019] and exploring alternative connectivity measures that could prove more robust than autoregressive amplitude-based approaches [Ruiz-Gómez et al., 2019].

Our proof-of-concept method is one of the first attempts to quantify connections between higher-order brain dynamics and cardiac outputs. This approach holds significant potential for comprehending large-scale neural functions and, in the context of PD, may serve as a tool for evaluating the effectiveness of dopamine treatments. The interactions between brain connectivity and cardiac dynamics can help us to better understand the complex physiopathology of PD, even in the early stages of the disease.

In conclusion, the investigation of large-scale neural dynamics within frameworks that integrate the interplay between higher-order brain dynamics and those occurring in peripheral organs presents a promising avenue for characterizing various diseases, extending beyond PD to encompass a range of neurodegenerative and neural damage conditions. This approach could not only broaden our understanding of the pathophysiology of such diseases but also paves the way for exploring cognitive states believed to involve higher-order dynamics. By conceiving brain functioning within an environment where internal organs play a significant role, this framework contributes to a paradigm shift that underscores the interconnected nature of brain function and peripheral physiology. Our framework opens new possibilities for comprehensive insights into the intricate relationships underpinning health and disease at both neural and systemic levels.

## Data Availability

The data is part of a publicly available dataset UC San Diego Resting State EEG Data from Patients with Parkinson's Disease, gathered from OpenNeuro.org the 21st of November of 2022

## Acknowledgements

The authors thank Alexander P. Rockhill, Nicko Jackson, Jobi George, Adam Aron, and Nicole C. Swann for sharing the data used in this study.

## Author contributions

Conceptualization: DCR

Methodology: DCR

Visualization: DCR

Investigation: DCR, MV, MC, FDVF

Supervision: MC, FDVF

Writing – first version of the manuscript: DCR

Writing – review & editing: DCR, MV, MC, FDVF

